# The effect of genetically proxied IL-6 signalling on severe malaria: A Mendelian randomisation analysis

**DOI:** 10.1101/2022.11.04.22281934

**Authors:** Fergus Hamilton, Ruth Mitchell, Andrei Constantinescu, David Hughes, Aubrey Cunnington, Peter Ghazal, Nicholas J Timpson

## Abstract

**Introduction:** Severe malaria remains a deadly disease for many young children in low- and middle-income countries. Levels of Interleukin-6 (IL-6) have been shown to identify cases of severe malaria and associate with severity, but it is unknown if this association is causal, or whether manipulation of IL-6 might alter outcomes in severe malaria.

**Methods:** A single nucleotide polymorphism (SNP, rs2228145) in the IL-6 receptor (*IL6R)* was chosen as a genetic variant that is known to alter IL-6 signalling. We measured the association between the minor allele of this SNP (C) and C-reactive protein (CRP) levels, a marker of IL-6 signalling in the non-European ancestry population recruited to UK Biobank.

We then took this forward as an instrument to perform Mendelian randomisation (MR) in MalariaGEN, a large cohort study of patients with severe malaria at eleven worldwide sites. As a secondary approach, we identified *cis* protein quantitative trait loci (*cis*-pQTL) for *IL6R* itself and other markers of IL-6 signalling in a recently published GWAS of the plasma proteome performed in African Americans. We then performed MR using these instruments in the African MalariaGEN sites (9/11). Analyses were performed at each site, and meta-analysed using inverse variance weighting. Additional analyses were performed for specific sub-phenotypes of severe malaria: cerebral malaria and severe malarial anaemia.

**Results:** The minor allele (C) of rs2228145 was associated with decreased CRP across all tested continental ancestries in UK Biobank. There was no evidence of heterogeneity of effect and a large overall effect (beta -0.11 per standard deviation of normalised CRP per C allele, p = 7.55 × 10^−255^)

In Mendelian randomisation studies using this SNP, we did not identify an effect of decreased IL-6 signalling on severe malaria case status (Odds ratio 1.14, 95% CI 0.56 – 2.34, p = 0.713). Estimates of the association with any severe malaria sub-phenotype were similarly null although there was significant imprecision in all estimates.

Using an alternative instrument (*cis*-pQTLs for *IL6R*), which included 3 SNPS (including rs2228145), we identified the same null effect, but with greater precision (Odds ratio 1.02, 95% CI 0.95 – 1.10), and no effect on any severe malaria subtypes.

**Conclusions:** Mendelian randomisation analyses using a SNP in the IL-6 receptor known to alter IL-6 signalling do not support a causal role for IL-6 signalling in the development of severe malaria, or any severe malaria sub-phenotype. This result suggests IL-6 may not be causal for severe outcomes in malaria, and that therapeutic manipulation of IL-6 may not be a suitable treatment for severe malaria.

## Introduction

Interleukin-6 (IL-6) is a critical cytokine in the innate immune response.^1^ Increased IL-6 is known to be associated with mortality within severe malaria, and studies have suggested that it may be causal for worse outcomes.^2,3^ However, animal models have so far failed to identify robust evidence that IL-6 manipulation (e.g. therapeutic blockade) improves outcomes in severe malaria, although data are inconclusive.^4^

In contrast, in other severe infections such as COVID-19, randomised trials have suggested that IL-6 is critical for pathogenesis, with evidence that blockade of IL-6 improves survival.^5^ In the large, recent WHO meta-analysis, IL-6 receptor antagonists (such as Tocilizumab) were associated with a 3-5% absolute decrease in mortality in patients critically ill with severe COVID-19.^5^ This trial evidence was predated by genetic studies using instrumental variable approaches (Mendelian randomisation^6^) that identified carriers of certain single nucleotide polymorphisms (SNPs) in the IL-6 receptor gene (*IL6R)* were relatively protected from severe COVID-19, a finding replicated in multiple independent cohorts.^7–10^ Recent similar studies also identified the same protective effect in sepsis, where variants in *IL6R* were again found to be protective against the development of sepsis, admission to critical care with sepsis, and death with sepsis.^8^

Given the trial evidence of effectiveness in COVID-19, and supporting genetic evidence in sepsis, we aimed to interrogate whether IL-6 downregulation might also lead to improved outcomes in severe malaria, and whether this might represent a common target for severe infection. The major challenge in undertaking similar two sample Mendelian randomisation (MR) analyses in malaria is the differing ancestries included in genetic studies of malaria as compared to those used to identify exposures.

Mendelian randomisation is a form of instrumental variable analysis whereby genetic variation (SNPs) known to associate with an exposure (in this case, IL-6 signalling) is used to estimate the effect of that exposure on an outcome (in this case, malaria), and under certain assumptions, can provide causal estimates.^6^ However, this relies on SNPs having the same effect on the exposure in the outcome dataset. As nearly all (>90%) of genome-wide association studies (GWAS) have been performed in people of European ancestry, while malaria genetic data is exclusively in non-European populations and linkage disequilibrium patterns and allele frequencies vary widely across genetic background, this poses a significant challenge to identifying SNPs that associate with an exposure and can be used in MR.^11^

In this study, we aimed to first identify SNPs that alter IL-6 signalling across non-European populations. Subsequently, we then used these SNPs in MR analyses to estimate the causal effect of IL-6 signalling on severe malaria.

## Methods

### Identification of exposures

We took two approaches to identifying exposures associated with IL-6 signalling to be used in MR. Firstly we used GWAS for C-reactive protein (CRP) in UK Biobank, of which there are substantial (n ∼ 19,000) non-European ancestry participants and identified variants that altered CRP but that were *cis* to *IL6R*. CRP is produced in hepatocytes in response to IL-6^1^, and is assumed to be a proxy for IL-6 signalling. This approach has been widely utilised in MR studies before.^7,8,12–14^

In European populations, the SNP rs2228145 (also known Asp358Ala) is known to alter levels of the IL-6 receptor and CRP, and has been shown to be functional in laboratory studies.^15,16^ In previous studies, this SNP explains between 20-40% of the variance in IL6R levels (and ∼1% of variance in CRP levels while multiple large scale genetic studies have implicated this SNP in a wide range of inflammatory and metabolic diseases.^17,18^ Studies using admixture mapping in admixed African American populations have also identified that this SNP as a causal candidate.^19^

We aimed to identify if rs2228145 was present and associated with CRP in populations outside Europe. Firstly, we extracted SNPs within 500kb up and downstream of the transcriptional start site of *IL6R* from previously performed GWAS of CRP across five non-European ancestry (EUR) groups (as defined by Pan-UKBB: African Ancestry (AFR), n = 6,203; Central and South Asian ancestry (CSA); n = 8,397; East Asian Ancestry (EAS), n = 2,564; Admixed American ancestry (AMR), n = 937; and Middle Eastern Ancestry (MID), n = 1,498) within UK Biobank, performed by the Pan-UKBB. All GWAS are available via the IEU OpenGWAS website,^20^ with details on genetic pre-processing, quality control, definition of continental ancestry group, and GWAS methodology on the Pan-UKBB website.^21^ Briefly, CRP was inverse-rank normal transformed and GWAS performed in each ancestry separately using SAIGE, a linear-mixed model approach.^22^

Subsequently, we meta-analysed these five GWAS using METAL^23^, in order to identify any heterogeneity and to identify if rs2228145 or any other SNPs had reliable associations in a trans-ancestry analysis. After meta-analysis, all SNPs that had a genome wide association (p < 5 × 10^−8^) were LD clumped (r2 < 0.01) using a 1000 Genomes African reference panel to identify if there was more than one independent signal at this locus.^24^ These SNPs were taken forward into MR analyses.

To increase precision of our effect estimates, and as effect estimates were similar across ancestries, we then included the much larger European ancestry GWAS performed by the pan-UKBB (EUR, n = 400,094) in our meta-analysis to generate effect estimates and standard errors.^21^

### Secondary exposures

As a secondary exposure and attempt to interrogate other aspects of the IL-6 axis, we utilised a recent GWAS of plasma protein levels performed in an African ancestry population (ARIC study, n ∼ 1,500).^17^ We identified three proteins associated with IL-6 function (IL-6 itself, the IL-6 receptor (IL6R) and gp130, which binds to IL6R to perform signalling)^1^ and identified variants within 500kb of the transcription start site of each protein that had a genome wide significant association with the protein (p < 5 × 10^−8^) and then performed LD clumping (r2 < 0.01) with the 1000 Genomes African ancestry LD reference panel using the TwoSampleMR package.^20^ These variants were then weighted by their association with the protein and taken forward for MR.

### Outcomes

For measurement of SNP-outcome associations, we used the large, geographically diverse MalariaGEN study which includes 11 populations, nine of which are in Africa.^25^ Detailed inclusion criteria are with the original study, but briefly, this study recruited cases of severe malaria using the WHO definition (and severe malaria subtypes) with population control and performed a GWAS in each ancestry of severe malaria case status followed by a meta-analysis across all sites.^25^ For our primary exposure (CRP), we used all MalariaGEN populations, but as our secondary exposures were all *cis-* PQTLs from African ancestry populations, we limited our study to the nine African populations.

The CRP associated SNP rs2228145 was directly genotyped in MalariaGEN using the Illumina Infium Omni 2.5M chip and has an INFO score of 1 across all study populations.

### Mendelian randomisation and meta-analysis

Mendelian randomisation is a form of instrumental variable analysis, that under certain assumptions can provide causal estimates of the effect of an exposure on an outcome. These assumptions are that the genetic instruments are associated with the risk factor of interest, were independent of potential confounders, and could only affect the outcome through the risk factor and not through alternative pathways (that is, through pleiotropy).^6^

We performed Mendelian randomisation using the rs2228145 SNP as an instrument (for our primary exposure, CRP) and used the SNP-CRP exposure from the cross ancestry meta-analysis to generate exposure weights. MR estimates were generated using the Wald ratio, or via inverse variance weighting when there was more than one SNP for each pQTL for our secondary exposures. MR estimates were then meta-analysed across each study site in an inverse variance weighted meta-analysis.

MR estimates from our primary exposure (CRP) analysis are in rank-inverse normal transformed units of change in CRP. MR was performed for each of the three severe malaria sub-types (severe malarial anaemia, cerebral malaria, and “other” severe malaria), with meta-analysis across populations as described above. For sensitivity analyses, for our *cis*-PQTL exposure, we also performed meta-analyses using MR Egger and weighted median approaches.

Analyses were performed using the TwoSampleMR package^20^ and R version 4.0.4 (R Foundation for Statistical Computing, Vienna).

### Ethics

As this study used only publicly available data, no ethical approval was required. Details of ethical approval for the datasets used in this study are available with the original publications: UK Biobank,^26^ ARIC,^17^ and MalariaGEN.^25^

### Guidelines

This study is reported in line with the STROBE-MR guidance, which is available as a supplement (Supplement S1)

### Data availability

This study was performed using publicly available data. MalariaGEN summary statistics are available at the MalariaGEN website^27^, while Pan-UKBB GWAS are available via the Pan-UKBB website^21^ and via the MRC-IEU OpenGWAS website.^20^

## Results

### Identification and assessment of variants at IL6R and association with CRP across ancestries

Across the six ancestry groups tested (European, Middle Eastern, African, Central South Asian, Admixed American, and East Asian), rs2228145 was associated with CRP levels, with little evidence of heterogeneity of effect (Table 1, p value for heterogeneity 0.67). The summary beta was -0.11 (SE = 0.012, p = 3.4 × 10^−21^) for each additional C allele excluding the European ancestry sub-population, and -0.11 (SE = 0.003, p = 7.55 × 10^−255^) including the much larger European ancestry sub-population.

**Table 1:** Effect of the rs2228145 allele on CRP across multiple continental ancestries in each of the Pan-UKB continental ancestry groups. CRP was inverse-rank normal transformed and so betas reflect a one SD change in inverse rank normal transformed CRP. The minor allele frequency (MAF) is from UK Biobank.

| Ancestry | Beta | SE | P value | MAF | n |
| --- | --- | --- | --- | --- | --- |
| *Individual ancestries:* |  |  |  |  |  |
| African<br>(AFR) | -0.111 | 0.03154 | $4.3 \times 10^{-4}$ | 0.098 | 6,203 |
| Admixed American<br>(AMR) | -0.134 | 0.04838 | $5.3 \times 10^{-3}$ | 0.450 | 937 |
| Central South Asian<br>(CSA) | -0.101 | 0.01565 | $8.7 \times 10^{-11}$ | 0.311 | 8,397 |
| East Asian<br>(EAS) | -0.144 | 0.02910 | $7.1 \times 10^{-7}$ | 0.342 | 2,564 |
| European<br>(EUR) | -0.106 | 0.00260 | $1.4 \times 10^{-320}$ | 0.410 | 400,094 |
| Middle Eastern<br>(MID) | -0.087 | 0.03549 | $1.35 \times 10^{-02}$ | 0.358 | 1,498 |
| *Meta-analysis* |  |  |  |  |  |
| Without European | -0.110 | 0.0116 | $3.37 \times 10^{-21}$ | | 19,599 |
| With European | -0.106 | 0.0031 | $7.55 \times 10^{-320}$ | | 419,693 |

The smallest beta was on the Middle Eastern ancestry population (beta -0.087) and the largest in the East Asian ancestry (beta -0.144), with study specific and meta-analysed results available in Table 1. The minor allele frequency was similar in all populations outside of Africa (0.30-0.40) but was much lower in the African ancestry group (0.098). Locus plots of this region are available in Figure 1.

**Figure 1:**
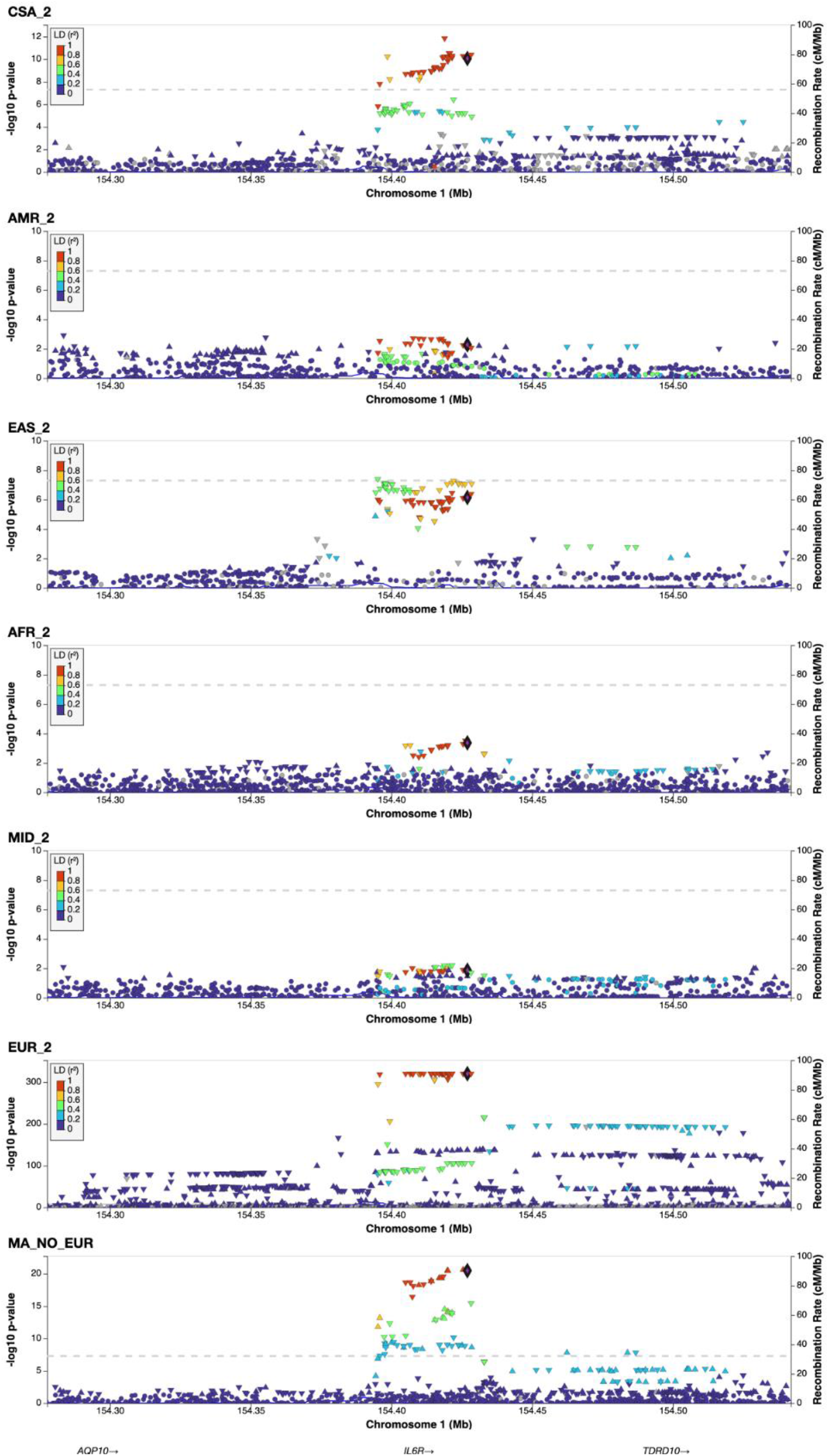
LocusZoom plots of the IL6R locus across each tested ancestry. LD values from the closest 1000 Genomes reference population. rs2228145 is the reference allele across all images.

We did not identify any other independent (r2 <0.01) variants within 500kb of the TSS of *IL6R* that had genome wide significance for CRP at this in the meta-analysed (excluding Europe) group. In summary, across all six continental ancestry groups, rs2228145 is associated with CRP (as a marker of IL-6 downregulation), has approximately similar effect size in all population, but the frequency of the C allele was lower in those of the African ancestry population.

### Identification of secondary exposures in the ARIC study

As secondary analyses we aimed to generate secondary instruments for other aspects of the IL-6 pathway. From the ARIC study^17^, we extracted GWAS for IL-6, IL6R, and gp130, and identified independent *cis*-PQTLs for each protein. We identified no cis pQTLs for IL-6, 3 for the IL6R (including rs2228145), and 1 for gp130. Included SNPs are listed in Table S1.

### Mendelian Randomisation

For our primary exposure (IL-6 signalling as measured by CRP) we performed MR using the sole variant rs2228145 as an instrument for CRP. Across all populations, we saw little evidence of any effect of IL-6 signalling on severe malaria, with all estimates crossing the null but with significant imprecision (Figure 2). In meta-analysis, our summary result was close to the null with an odds ratio (OR) of 1.21 (95% CI 0.51 – 2.88 p = 0.67) per each normalized unit increase in CRP. Figure 2 shows the results of this analysis, with each individual population and the summary random-effects meta-analysis result, with number of cases at each site, and study site effect estimates in Table 2. We did not identify evidence of study site specific effects (I2 = 0, p for heterogeneity = 0.532).

**Table 2:** MR Effect estimates for each of the 11 populations, generated by a Wald ratio, and number of cases at each site. MR estimates are on the scale of a one SD increase in inverse rank normal transformed CRP.

| Country | Odds ratio | P value | Total number | Cases (% total) | SMA (% cases) | CM (% cases) |
| --- | --- | --- | --- | --- | --- | --- |
| Burkina Faso | 9.6 (0.32 - 291.82) | 0.194 | 1327 | 733 (55.2%) | 28 (3.8%) | 94 (12.8%) |
| Cameroon | 0.16 (0.01 - 3.81) | 0.260 | 1277 | 592 (46.4%) | 66 (11.1%) | 32 (5.4%) |
| Gambia | 1.54 (0.43 - 5.6) | 0.509 | 5091 | 2487 (48.9%) | 456 (18.3%) | 780 (31.4%) |
| Ghana | 3.83<br>(0.09 -<br>165.93) | 0.485 | 716 | 396 (55.3%) | 41 (10.4%) | 31 (7.8%) |
| Kenya | 0.38<br>(0.07 -<br>2.05) | 0.261 | 3261 | 1646 (50.5%) | 174 (10.6%) | 690 (41.9%) |
| Malawi | 0.78<br>(0.09 -<br>7.07) | 0.823 | 2499 | 1182 (47.3%) | 65 (5.5%) | 642 (54.3%) |
| Mali | 0.17 (0 -<br>20.51) | 0.467 | 446 | 263 (59%) | 81 (30.8%) | 61 (23.2%) |
| Nigeria | 0 (0 -<br>822.19) | 0.295 | 131 | 109 (83.2%) | 1 (0.9%) | 28 (25.7%) |
| PNG | 1.11<br>(0.04 -<br>27.77) | 0.947 | 770 | 396 (51.4%) | 115 (29%) | 49 (12.4%) |
| Tanzania | 10.91<br>(0.36 -<br>333.18) | 0.171 | 807 | 409 (50.7%) | 178 (43.5%) | 31 (7.6%) |
| Vietnam | 1.86 (0.3<br>- 11.57) | 0.507 | 1264 | 718 (56.8%) | 23 (3.2%) | 154 (21.4%) |
| IVW: Meta-analysis<br>(random<br>effects) | 1.21<br>(0.51 -<br>2.88) | 0.670 | 17,589 | 8,931 | 1,228 | 2,592 |

**Figure 2:**
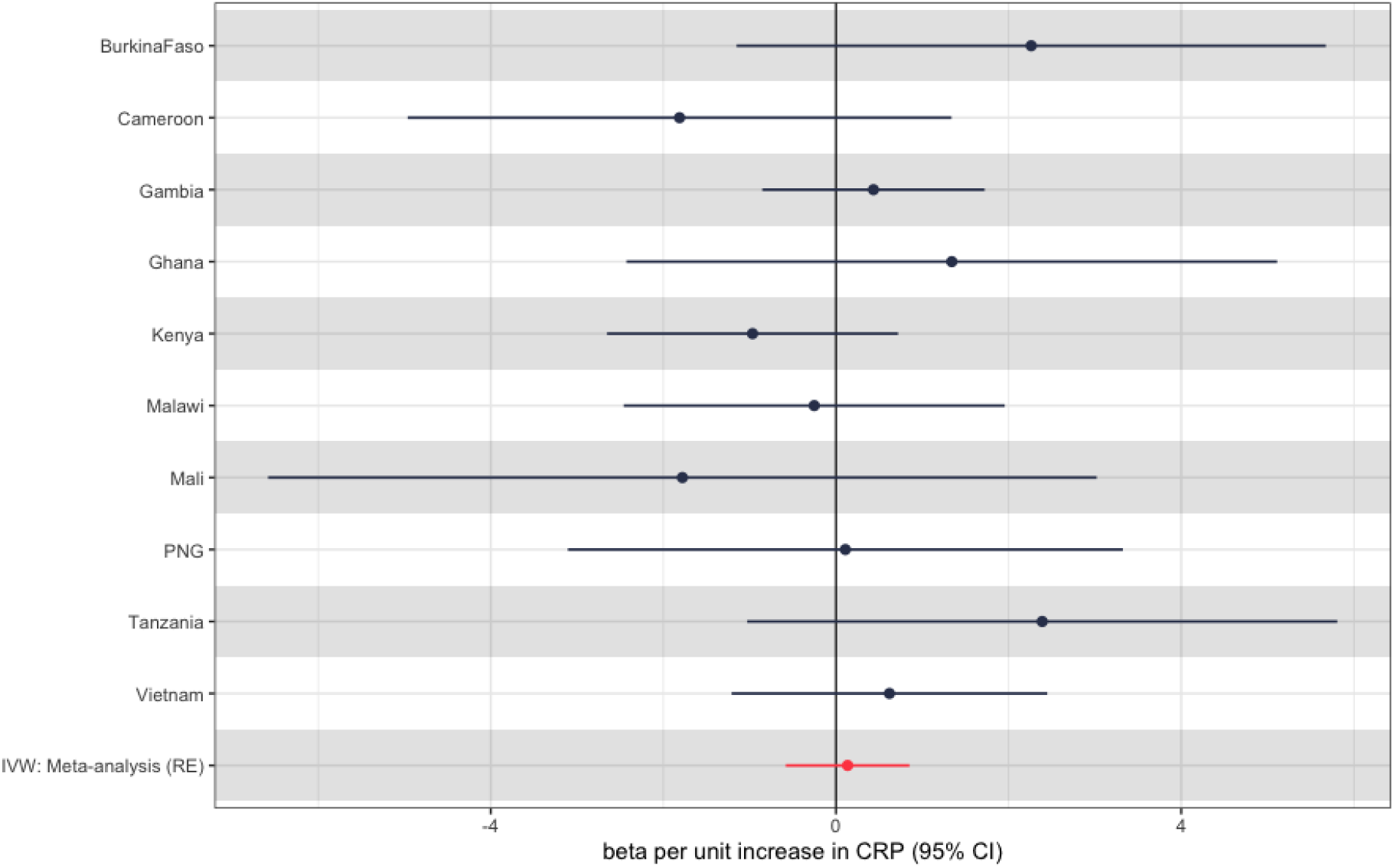
MR effecte estimates for each population and the summary inverse variance weighted beta for the rs2228145 SNP. Note, Nigeria not shown due to presence of a single case of severe malaria (Table 2). Effect estimates generated by the Wald Ratio.

We then went on to perform MR for three malaria subtypes. As with the main analysis, these results were largely null, but were imprecise and did not preclude small effects. Figure 3 shows these effects for each sub-phenotype of severe malaria, with Table S2 showing the raw estimates.

**Figure 3:**
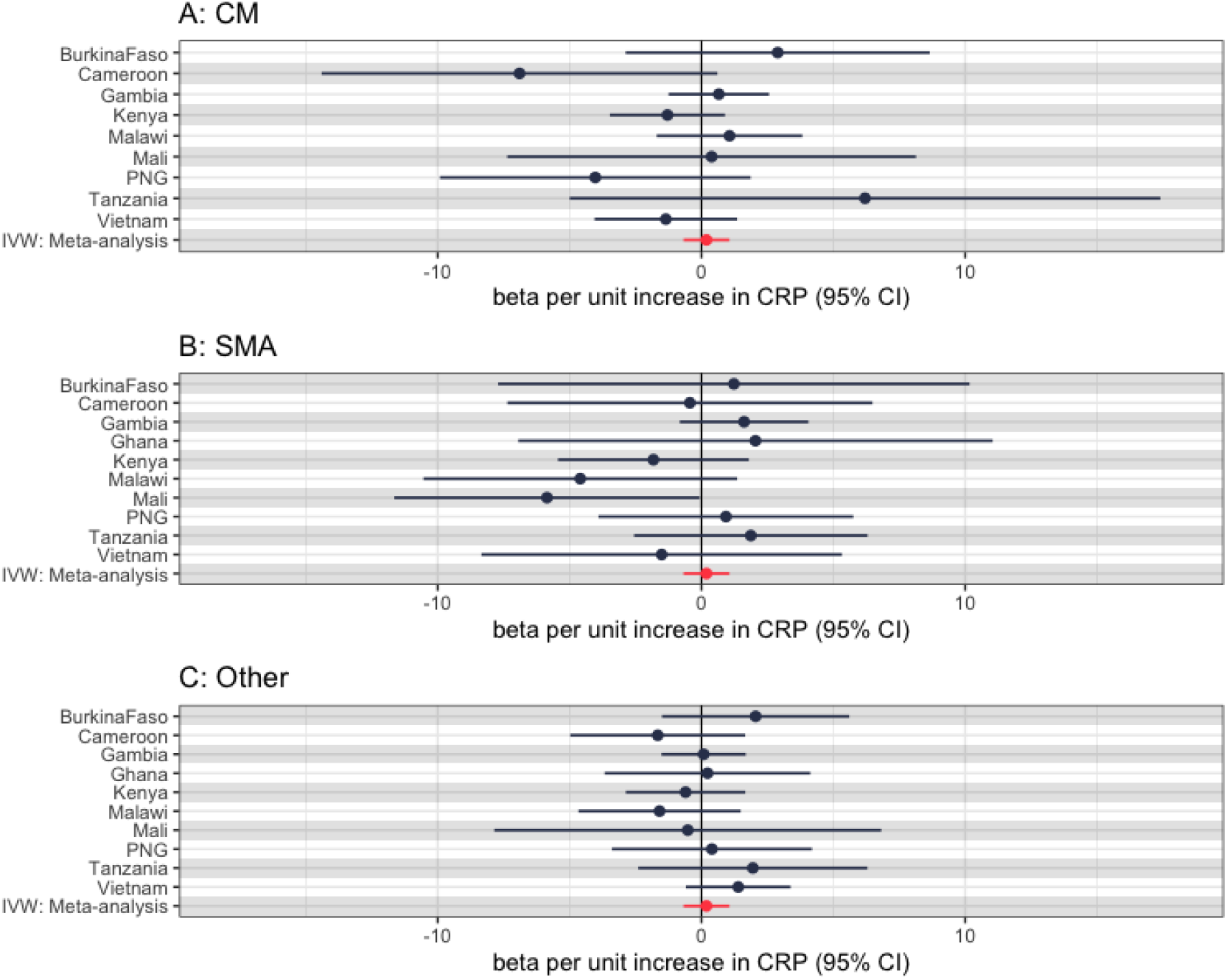
MR effect estimates (via the Wald ratio) for each severe malaria subtype using the rs2228145 SNP, with the summary IVW meta-analysis effect estimate also (A: Cerebral Malaria (CM), B: Severe Malarial Anaemia (SMA), C: Other). Note, Nigeria not shown again for all subtypes due to low case numbers, Ghana not shown for CM subtype only due to low case numbers.

### Secondary exposures

We then went on to perform MR using *cis*-PQTLs for *gp130* and *IL6R* generated from the ARIC study^17^, which was undertaken in an African American population. At the *IL6R* locus, we identified three *cis*-PQTLs (one of which was rs2228145), enabling us to perform inverse-variance weighted meta-analysis and increase our power. Analyses yielded a summary MR estimate of an odds ratio of 1.02 (95% CI 0.95 – 1.10) per each SD increase in inverse-rank normalised IL6R protein levels, with confidence intervals for all study sites crossing the null (Figure 4). Alternative meta-analysis methods (MR Egger and Weighted median) are reported in Table S3 but had very similar results.

**Figure 4:**
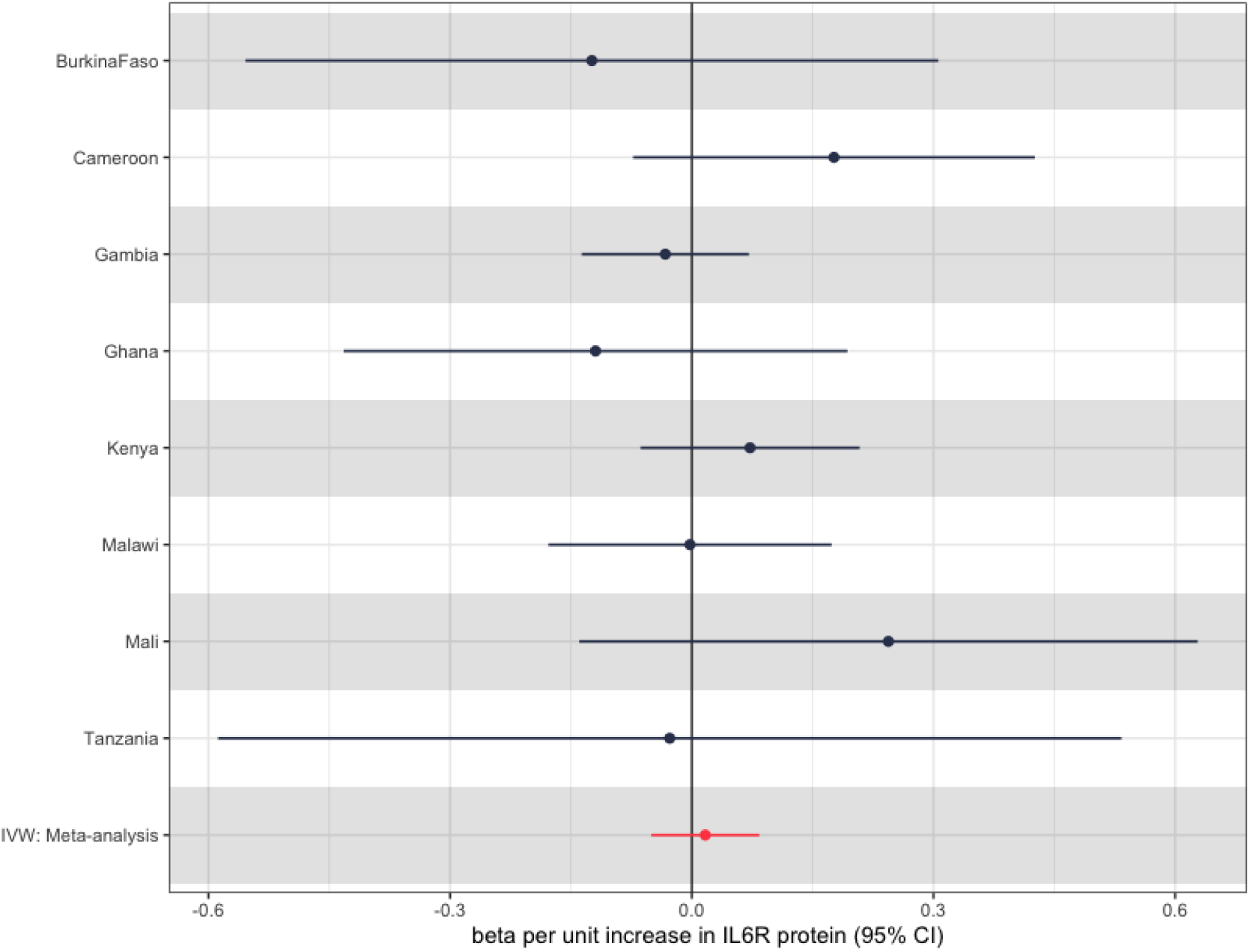
Inverse variance weighted MR estimates for each study site for the association between *IL6R* protein levels and severe malaria case status. These are on the scale of an SD increase in inverse-rank normalised transformed IL6R protein levels. Note Nigeria again not shown due to low numbers of cases.

When looking at the sub-phenotypes of malaria, we saw a similar null result, with again a degree of imprecision due to low case numbers of each subtype at certain sites. Figure 5 shows this, with results shown in Table S4

**Figure 5:**
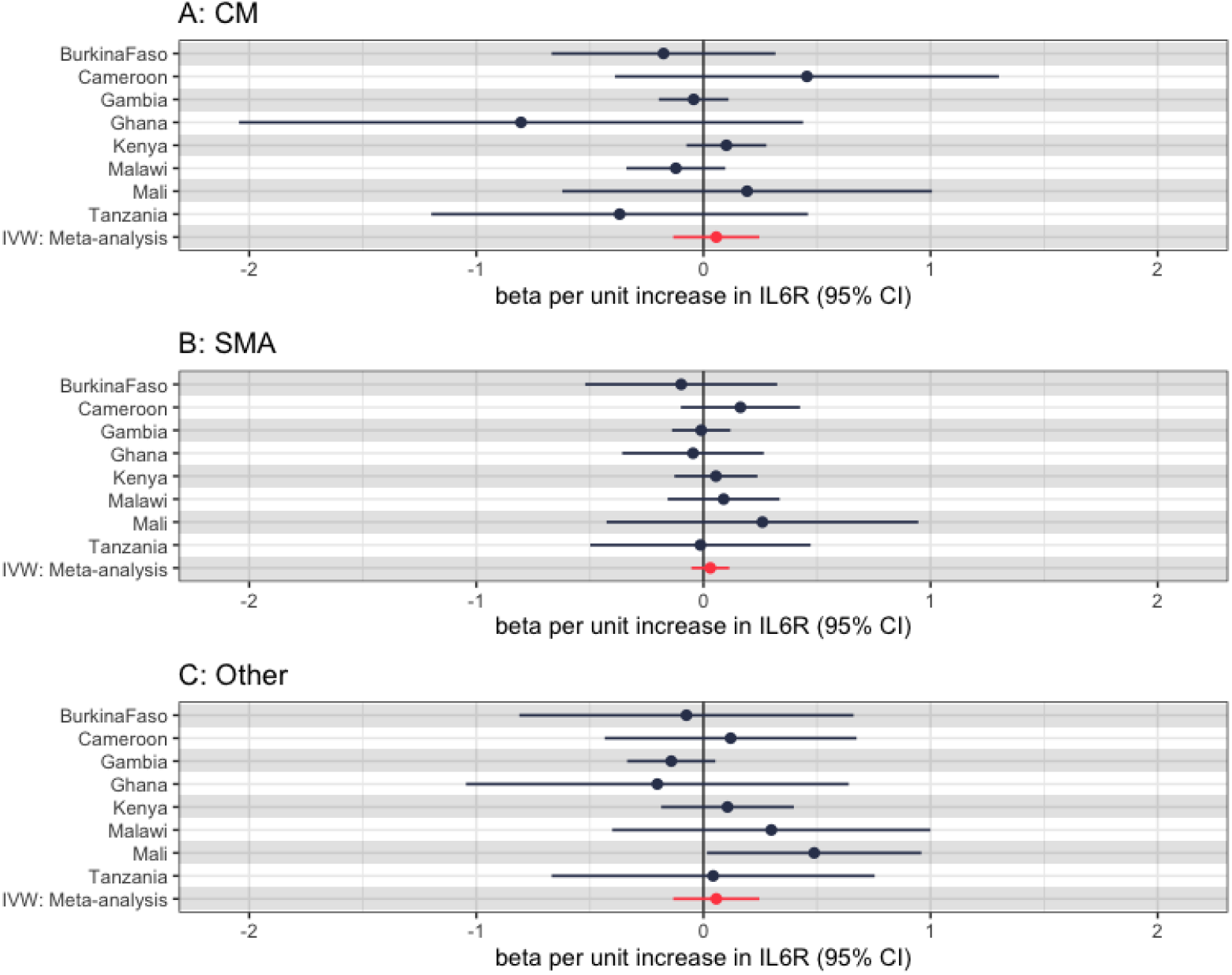
MR effect estimates (via the Wald ratio) for each severe malaria subtype (A: Cerebral Malaria (CM), B: Severe Malarial Anaemia (SMA), C: Other). Note, Nigeria not shown again due to significant imprecision.

### gp130

*gp130* encodes for the protein gp130 (also known as IL6ST), which is the other subunit of the IL-6 receptor. We identified one *cis*-pQTL for this and performed MR using the Wald ratio to generate estimate. Again, we identified a null effect, with a summary OR of 1.06 (0.88 - 1.28) for each SD increase in gp130 (Figure S1), with similar results for each malaria subtype (Figure S2). Results for the main analysis are shown in Table S5, with results for the subtype analyses in Table S6

## Conclusions

In this study, we investigated whether SNPs, that were selected on the basis of interrupting IL6R and assumed to be a proxy for IL-6 signalling, are associated with severe malaria case status.

Firstly, we showed that the rs2228145 variant in *IL6R* - a known splicing variant - associates with CRP levels (a marker of IL-6 signalling) across all tested ancestries in UK Biobank and explains around 40-60% of variance in IL6R levels in people of African ancestry^17^, and therefore represents a good instrument to perform MR across non-European ancestries. We then took this forward in MR analyses. We could not identify any effect of rs2228145 on severe malaria case status, or any severe malaria sub-phenotype, although estimates remained imprecise. However, when subsequently using multiple *cis*-pQTLs for *IL6R*, which are known to alter downstream IL-6 signalling we identified the same apparently null result with more precision in the African populations included in MalariaGEN^25^. Additionally, analyses undertaken did not identify any association with other proteins in the IL-6 signalling pathway.

This work leaves several questions about the role of IL-6 in severe malaria. A recent meta-analysis confirmed that IL-6 is prognostic for severe outcomes, and differentiates uncomplicated from severe malaria.^2^ However, our work suggests, in contrast to COVID-19 (and perhaps bacterial sepsis), those with genetically proxied reduced IL-6 signalling do not have altered risk of severe disease.^7,8^ IL-6 levels are raised (and associate with) a huge number of conditions, and so it is plausible that IL-6 may represent a useful biomarker but one that is not causally related to severe malaria. Supporting this, the results of animal models of IL-6 manipulation in malaria have been inconsistent.^3,4^ In summary, our work supports the hypothesis that IL-6 is not causal for severe malaria.

### Limitations

Like many Mendelian randomisation studies, this study is limited by the available exposure and outcome data, and the assumptions of Mendelian randomisation. As all populations in MalariaGEN are outside Europe, and there are few non-European large scale GWAS of inflammatory biomarkers, we are limited to utilising exposure data from the non-European population of UK Biobank (n ∼ 19,000)^28^, and the recently published ARIC study (n ∼ 1,500).^17^ As far as we are aware, there are no other available sources of potential data in non-European populations. For contrast, the recent GWAS of C-reactive protein in European ancestry populations included 557,000 people.^29^

Because of this, for our main exposure, we were limited to using the well understood rs2228145 SNP, which has been widely used for MR in European ancestry populations.^8,12,30^ Although *in-vitro* work has confirmed this variant reduces cell surface expression of IL6R,^16^ and large scale transcriptome studies in healthy patients^31^ and those with infection^32^ have shown this is a splicing variant, the use of a single variant reduces the statistical power of the study. This is compounded by the low minor allele frequency (∼10%) in African ancestry populations, reducing the power to identify small associations further. In order to overcome this, our secondary exposure used multiple cis-pQTLs for IL6R, a key IL-6 signalling molecule, and identified the same null result but with much greater precision, supporting the lack of effect at this locus.

The assumptions of MR (relevance, independence, and exclusion restriction) are largely unfalsifiable. However, given the location of our primary exposure (*cis* to the gene of interest), the known biological function of the IL-6 receptor, and the extensive literature supporting MR at this locus,^7,13,33,34^, including in severe infection,^10^ we can be more confident of our results.

Our final limitation remains the challenge of interpreting germline variation relating to a lifetime exposure to changes in IL-6 signalling as evidence for or against the therapeutic usage of IL-6 antagonism in acute malaria, or for elucidating the causal role of IL-6 in severe malaria. This evidence is suggestive that IL-6 is not causal, and should be taken in the context that variants that alter IL-6 signalling do alter both the incidence and outcomes of other infections, and that trial evidence of IL-6 inhibition has supported the genetic evidence in COVID-19.^7,8,35^ However, we would caution over-interpretation of our null result to suggest IL-6 is irrelevant in severe malaria, although it does weaken the case for suggestion of IL-6 inhibition as a therapeutic option.

### Conclusions

Using SNPs near *IL6R* to proxy IL-6 signalling, we found no evidence that IL-6 signalling has a causal role in the development of severe malaria, although our results had imprecision that cannot preclude a small effect. This evidence does not support the consideration of IL-6 manipulation in patients with severe malaria.

## Supporting information

STROBE MR

## Data Availability

This study was performed using publicly available data. MalariaGEN summary statistics are available at the MalariaGEN website 27 while Pan-UKBB GWAS are available via the Pan-UKBB website 21 and via the MRC-IEU OpenGWAS website.20

## Funding, support and the role of the funding source

FHs time was funded by the GW4-CAT Wellcome Doctoral Fellowship Scheme. PG’s time was funded by the Ser Cymru programme, the Welsh Government, and the EU-ERDF.

NJT is a Wellcome Trust Investigator (202802/Z/16/Z), is the PI of the Avon Longitudinal Study of Parents and Children (MRC & WT 217065/Z/19/Z), is supported by the University of Bristol NIHR Biomedical Research Centre (BRC-1215-2001), the MRC Integrative Epidemiology Unit (MC_UU_00011/1) and works within the CRUK Integrative Cancer Epidemiology Programme (C18281/A29019). This study makes use of data generated by MalariaGEN. A full list of the investigators who contributed to the generation of the data is available from www.malariagen.net. Funding for this project was provided by Wellcome Trust (WT077383/Z/05/Z) and the Bill & Melinda Gates Foundation through the Foundation of the National Institutes of Health (566) as part of the Grand Challenges in Global Health Initiative. The funder had no role in the design, analysis, or reporting of this study.

## Conflicts of interest

No authors declare any relevant conflicts of interest.

## Notes

### Competing Interest Statement

The authors have declared no competing interest.

### Funding Statement

FHs time was funded by the GW4-CAT Wellcome Doctoral Fellowship Scheme. PGs time was funded by the Ser Cymru programme, the Welsh Government, and the EU-ERDF. 
NJT is a Wellcome Trust Investigator (202802/Z/16/Z), is the PI of the Avon Longitudinal Study of Parents and Children (MRC & WT 217065/Z/19/Z), is supported by the University of Bristol NIHR Biomedical Research Centre (BRC-1215-2001), the MRC Integrative Epidemiology Unit (MC_UU_00011/1) and works within the CRUK Integrative Cancer Epidemiology Programme (C18281/A29019). This study makes use of data generated by MalariaGEN. A full list of the investigators who contributed to the generation of the data is available from www.malariagen.net. Funding for this project was provided by Wellcome Trust (WT077383/Z/05/Z) and the Bill & Melinda Gates Foundation through the Foundation of the National Institutes of Health (566) as part of the Grand Challenges in Global Health Initiative. The funder had no role in the design, analysis, or reporting of this study.

### Author Declarations

This study only used publicly available summary genetic association data.

