## Supplementary material for "The effect of genetically proxied IL-6 signalling on severe malaria: A Mendelian randomisation analysis": Supp Figs

Supplementary Figures:

**Figure S1**: Raw inverse variance weighted MR estimates for each study site for the association between gp130 protein levels and severe malaria case status (via Wald Ratio). These are on the scale of an SD increase in inverse-rank normalised transformed gp130 protein levels. Note Nigeria again not shown due to imprecision.

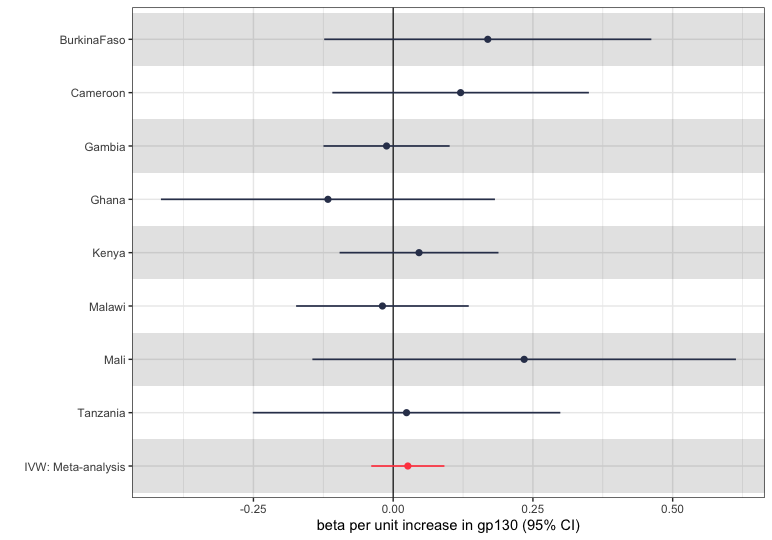

**Figure S2**: Raw inverse variance weighted MR estimates for each study site for the association between gp130 protein levels and severe malaria subtype status (via Wald ratio). These are on the scale of an SD increase in inverse-rank normalised transformed gp130 protein levels. Note Nigeria again not shown due to imprecision.

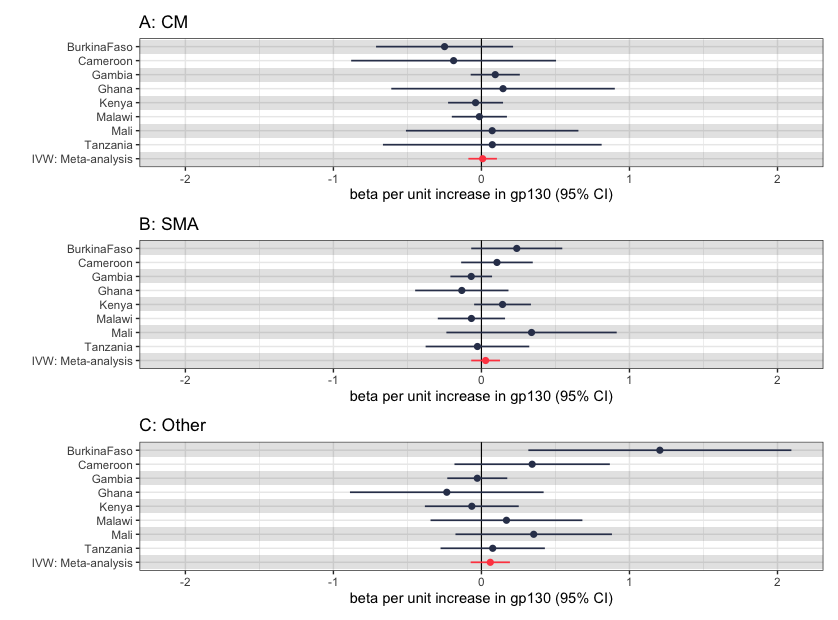
