## Supplementary material for "The effect of genetically proxied IL-6 signalling on severe malaria: A Mendelian randomisation analysis": STROBE MR

**STROBE-MR checklist of recommended items to address in reports of Mendelian randomization studies**^1^ ^2^

| **Item No.** | **Section** | **Checklist item** | **Page No.** | **Relevant text from manuscript** |
| --- | --- | --- | --- | --- |
| 1 | **TITLE and ABSTRACT** | Indicate Mendelian randomization (MR) as the study’s design in the title and/or the abstract if that is a main purpose of the study | 1 | The effect of genetically proxied IL-6 signalling on severe malaria: A Mendelian randomisation analysis |
|  | **INTRODUCTION** |  |  |  |
| 2 | **Background** | Explain the scientific background and rationale for the reported study. What is the exposure? Is a potential causal relationship between exposure and outcome plausible? Justify why MR is a helpful method to address the study question | 3 | Mendelian randomisation is a form of instrumental variable analysis whereby genetic variation (SNPs) known to associate with an exposure (in this case, IL-6 signalling) is used to estimate the effect of that exposure on an outcome (in this case, malaria), and under certain assumptions, can provide causal estimates |
| 3 | **Objectives** | State specific objectives clearly, including pre-specified causal hypotheses (if any). State that MR is a method that, under specific assumptions, intends to estimate causal effects | 4 | Subsequently, we then used these SNPs in MR analyses to estimate the causal effect of IL-6 signalling on severe malaria. |
|  | **METHODS** |  |  |  |
| 4 | **Study design and data sources** | Present key elements of the study design early in the article. Consider including a table listing sources of data for all phases of the study. For each data source contributing to the analysis, describe the following: | 4 | Sections: Exposures, Outcomes |
|  | a) | Setting: Describe the study design and the underlying population, if possible. Describe the setting, locations, and relevant dates, including periods of recruitment, exposure, follow-up, and data collection, when available. | 4 | See above. |
|  | b) | Participants: Give the eligibility criteria, and the sources and methods of selection of participants. Report the sample size, and whether any power or sample size calculations were carried out prior to the main analysis | 4 | See above, and references. |
|  | c) | Describe measurement, quality control and selection of genetic variants | 4,6 | All GWAS are available via the IEU OpenGWAS website,^20^ with details on genetic pre-processing, quality control, definition of continental ancestry group, and GWAS methodology on the Pan-UKBB website.^21^ Briefly, CRP was inverse-rank normal transformed and GWAS performed in each ancestry separately using SAIGE, a linear-mixed model approach.^22^  The CRP associated SNP rs2228145 was directly genotyped in MalariaGEN using the Illumina Infium Omni 2.5M chip and has an INFO score of 1 across all study populations. |
|  | d) | For each exposure, outcome, and other relevant variables, describe methods of assessment and diagnostic criteria for diseases | 5 | Ref 22, page 5 |
|  | e) | Provide details of ethics committee approval and participant informed consent, if relevant |  | NR |
| 5 | **Assumptions** | Explicitly state the three core IV assumptions for the main analysis (relevance, independence and exclusion restriction) as well assumptions for any additional or sensitivity analysis | 6 | Mendelian randomisation is a form of instrumental variable analysis, that under certain assumptions can provide causal estimates of the effect of an exposure on an outcome. These assumptions are that the genetic instruments are associated with the risk factor of interest, were independent of potential confounders, and could only affect the outcome through the risk factor and not through alternative pathways (that is, through pleiotropy). |
| 6 | **Statistical methods: main analysis** | Describe statistical methods and statistics used | 6,7 | We performed Mendelian randomisation using the rs2228145 SNP as an instrument (for our primary exposure, CRP) and used the SNP-CRP exposure from the cross ancestry meta-analysis to generate exposure weights. MR estimates were generated using the Wald ratio, or via inverse variance weighting when there was more than one SNP for each pQTL for our secondary exposures. MR estimates were then meta-analysed across each study site in an inverse variance weighted meta-analysis.  MR estimates from our primary exposure (CRP) analysis are in rank-inverse normal transformed units of change in CRP. MR was performed for each of the three severe malaria sub-types (severe malarial anaemia, cerebral malaria, and “other” severe malaria), with meta-analysis across populations as described above. For sensitivity analyses, for our cisPQTL exposure, we also performed meta-analyses using MR Egger and weighted median approaches. |
|  | a) | Describe how quantitative variables were handled in the analyses (i.e., scale, units, model) | 4 | 4, |
|  | b) | Describe how genetic variants were handled in the analyses and, if applicable, how their weights were selected | 4-6 | Multiple text |
|  | c) | Describe the MR estimator (e.g. two-stage least squares, Wald ratio) and related statistics. Detail the included covariates and, in case of two-sample MR, whether the same covariate set was used for adjustment in the two samples | 6 | Wald Ratio, see above |
|  | d) | Explain how missing data were addressed | NR |  |
|  | e) | If applicable, indicate how multiple testing was addressed | NR |  |
| 7 | **Assessment of assumptions** | Describe any methods or prior knowledge used to assess the assumptions or justify their validity | 4-6, 17-18 | Discussion, Intro |
| 8 | **Sensitivity analyses and additional analyses** | Describe any sensitivity analyses or additional analyses performed (e.g. comparison of effect estimates from different approaches, independent replication, bias analytic techniques, validation of instruments, simulations) | 6-7 | MR Egger, Weighted Median |
| 9 | **Software and pre-registration** |  |  |  |
|  | a) | Name statistical software and package(s), including version and settings used | 6 |  |
|  | b) | State whether the study protocol and details were pre-registered (as well as when and where) | NR |  |
|  | **RESULTS** |  |  |  |
| 10 | **Descriptive data** |  |  |  |
|  | a) | Report the numbers of individuals at each stage of included studies and reasons for exclusion. Consider use of a flow diagram | 7 | Table 1 |
|  | b) | Report summary statistics for phenotypic exposure(s), outcome(s), and other relevant variables (e.g. means, SDs, proportions) | 7 | Table 1 |
|  | c) | If the data sources include meta-analyses of previous studies, provide the assessments of heterogeneity across these studies | 7 | Table 1, text |
|  | d) | For two-sample MR:  i.  Provide justification of the similarity of the genetic variant-exposure associations between the exposure and outcome samples  ii.  Provide information on the number of individuals who overlap between the exposure and outcome studies | 4-6 | Introduction, ancestry. Nil overlap. |
| 11 | **Main results** |  |  |  |
|  | a) | Report the associations between genetic variant and exposure, and between genetic variant and outcome, preferably on an interpretable scale | 8 | Table 2, Figure 1, Table S2 |
|  | b) | Report MR estimates of the relationship between exposure and outcome, and the measures of uncertainty from the MR analysis, on an interpretable scale, such as odds ratio or relative risk per SD difference | 8 | Table 2, Figure 1, Table S2 |
|  | c) | If relevant, consider translating estimates of relative risk into absolute risk for a meaningful time period | NR |  |
|  | d) | Consider plots to visualize results (e.g. forest plot, scatterplot of associations between genetic variants and outcome versus between genetic variants and exposure) | 8 | Table 2, Figure 1, Table S2 |
| 12 | **Assessment of assumptions** |  |  |  |
|  | a) | Report the assessment of the validity of the assumptions | 16,17 | Like many Mendelian randomisation studies, this study is limited by the available exposure and outcome data, and the assumptions of Mendelian randomisation. As all populations in MalariaGEN are outside Europe, and there are few non-European large scale GWAS of inflammatory biomarkers, we are limited to utilising exposure data from the non-European population of UK Biobank (n ~ 19,000)^27^, and the recently published ARIC study (n ~ 1,500).^17^ As far as we are aware, there are no other available sources of potential data in non-European populations. For contrast, the recent GWAS of C-reactive protein in European ancestry populations included 557,000 people.^28^ |
|  | b) | Report any additional statistics (e.g., assessments of heterogeneity across genetic variants, such as *I^2^*, Q statistic or E-value) |  | NR |
| 13 | **Sensitivity analyses and additional analyses** |  |  |  |
|  | a) | Report any sensitivity analyses to assess the robustness of the main results to violations of the assumptions | 12-14 |  |
|  | b) | Report results from other sensitivity analyses or additional analyses | 12-14 |  |
|  | c) | Report any assessment of direction of causal relationship (e.g., bidirectional MR) | NR |  |
|  | d) | When relevant, report and compare with estimates from non-MR analyses | NR |  |
|  | e) | Consider additional plots to visualize results (e.g., leave-one-out analyses) | NR |  |
|  | **DISCUSSION** |  |  |  |
| 14 | **Key results** | Summarize key results with reference to study objectives | 16 |  |
| 15 | **Limitations** | Discuss limitations of the study, taking into account the validity of the IV assumptions, other sources of potential bias, and imprecision. Discuss both direction and magnitude of any potential bias and any efforts to address them | 16-18 |  |
| 16 | **Interpretation** |  |  |  |
|  | a) | Meaning: Give a cautious overall interpretation of results in the context of their limitations and in comparison with other studies | 16-18 |  |
|  | b) | Mechanism: Discuss underlying biological mechanisms that could drive a potential causal relationship between the investigated exposure and the outcome, and whether the gene-environment equivalence assumption is reasonable. Use causal language carefully, clarifying that IV estimates may provide causal effects only under certain assumptions | 16 |  |
|  | c) | Clinical relevance: Discuss whether the results have clinical or public policy relevance, and to what extent they inform effect sizes of possible interventions | 16-18 |  |
| 17 | **Generalizability** | Discuss the generalizability of the study results (a) to other populations, (b) across other exposure periods/timings, and (c) across other levels of exposure | 16-18 |  |
|  | **OTHER INFORMATION** |  |  |  |
| 18 | **Funding** | Describe sources of funding and the role of funders in the present study and, if applicable, sources of funding for the databases and original study or studies on which the present study is based | 18 | FH’s time was funded by the GW4-CAT Wellcome Doctoral Fellowship Scheme. PG’s time was funded by the Ser Cymru programme, the Welsh Government, and the EU-ERDF.  NJT is a Wellcome Trust Investigator (202802/Z/16/Z), is the PI of the Avon Longitudinal Study of Parents and Children (MRC & WT 217065/Z/19/Z), is supported by the University of Bristol NIHR Biomedical Research Centre (BRC-1215-2001), the MRC Integrative Epidemiology Unit (MC_UU_00011/1) and works within the CRUK Integrative Cancer Epidemiology Programme (C18281/A29019). This study makes use of data generated by MalariaGEN. A full list of the investigators who contributed to the generation of the data is available from www.malariagen.net. Funding for this project was provided by Wellcome Trust (WT077383/Z/05/Z) and the Bill & Melinda Gates Foundation through the Foundation of the National Institutes of Health (566) as part of the Grand Challenges in Global Health Initiative. The funder had no role in the design, analysis, or reporting of this study. |
| 19 | **Data and data sharing** | Provide the data used to perform all analyses or report where and how the data can be accessed, and reference these sources in the article. Provide the statistical code needed to reproduce the results in the article, or report whether the code is publicly accessible and if so, where | 7 | This study was performed using publicly available data. MalariaGEN summary statistics are available at the MalariaGEN website^27^, while Pan-UKBB GWAS are available via the Pan-UKBB website^21^ and via the MRC-IEU OpenGWAS website.^20^ |
| 20 | **Conflicts of Interest** | All authors should declare all potential conflicts of interest | 18 | No authors declare any relevant conflicts of interest. |

This checklist is copyrighted by the Equator Network under the Creative Commons Attribution 3.0 Unported (CC BY 3.0) license.

1.	Skrivankova VW, Richmond RC, Woolf BAR, Yarmolinsky J, Davies NM, Swanson SA, et al. Strengthening the Reporting of Observational Studies in Epidemiology using Mendelian Randomization (STROBE-MR) Statement. JAMA. 2021;under review.

2.	Skrivankova VW, Richmond RC, Woolf BAR, Davies NM, Swanson SA, VanderWeele TJ, et al. Strengthening the Reporting of Observational Studies in Epidemiology using Mendelian Randomisation (STROBE-MR): Explanation and Elaboration. BMJ. 2021;375:n2233.
